# Probabilistic Cerebral Blood Flow Trajectories Across the Adult Lifespan Using Quantitative [^15^O]Water PET

**DOI:** 10.64898/2026.04.08.26350393

**Authors:** Jarkko Johansson, Santeri Palonen, Ksenia Egorova, Jouni Tuisku, Harri Harju, Henri Kärpijoki, Teemu Maaniitty, Antti Saraste, Teemu Saari, Nelli Tuomola, Juha Rinne, Pirjo Nuutila, Aino Latva-Rasku, Kirsi A. Virtanen, Juhani Knuuti, Lauri Nummenmaa

## Abstract

**Background:** Quantitative cerebral blood flow (CBF) measured with [^15^O]water positron emission tomography (PET) is the reference standard for quantifying brain perfusion. However, clinical interpretation of individual CBF measurements is limited by the absence of large normative datasets accounting for physiological variability across the adult lifespan. Long–axial field-of-view PET enables high-sensitivity quantitative [^15^O]water perfusion imaging without arterial blood sampling, allowing normative characterization of cerebral perfusion at unprecedented scale. The aim of this study was to establish normative and covariate-adjusted models of cerebral blood flow across the adult lifespan using total-body [^15^O]water PET.

**Methods:** Quantitative CBF measurements were obtained in 302 neurologically healthy adults (age 21–86 years) using total-body [^15^O]water PET. Linear mixed-effects models were used to evaluate the effects of age, sex, body mass index (BMI), and blood hemoglobin concentration on CBF and to generate normative prediction models across the adult lifespan. Between-subject and within-subject variability were estimated from repeated scans in a subset of participants (n=51).

**Results:** Mean grey matter CBF was 46.1 mL/(min*dL), with substantial inter-individual variability but high within-subject reproducibility (intraclass correlation coefficients 0.78–0.89). Advancing age was associated with a decline in CBF of approximately 7% per decade (p_FDR < 10^-12^). Higher BMI was associated with lower CBF (approximately −6% per 10 kg/m^2^; p_FDR < 0.01). Women exhibited higher CBF than men (approximately 7.5%), but this difference was largely explained by lower blood hemoglobin concentration in women. Covariate-adjusted models were used to generate normative predictions and prediction intervals describing expected CBF across adulthood.

**Conclusion:** This study establishes a normative database of quantitative cerebral blood flow across the adult lifespan using high-sensitivity [^15^O]water PET. Age, BMI, and hemoglobin are major determinants of inter-individual variability in CBF. The resulting generative models provide a quantitative reference framework for interpreting cerebral perfusion measurements and may enable automated detection of abnormal brain perfusion in clinical PET imaging.

## Introduction

The brain receives approximately 15–20% of the cardiac output while accounting for roughly 20% of the body’s total oxygen consumption, despite representing only about 2% of body mass (*1*). Because the brain lacks substantial energy reserves, continuous delivery of oxygen and substrates is essential for neuronal function. Cerebral blood flow (CBF) is therefore tightly regulated through autoregulatory mechanisms that maintain relatively stable perfusion across a range of systemic blood pressures and metabolic demands (*2*). Disruption of cerebral perfusion rapidly leads to ischemia and irreversible neuronal injury, ultimately impairing cognitive and neurological function (*3*).

Cerebrovascular disease is among the leading causes of morbidity and mortality worldwide (*4*). While large-vessel occlusions typically manifest as clinically recognized stroke, a substantial proportion of cerebrovascular injury occurs silently (*5*). Cerebral small vessel disease (cSVD), one of the most common neurological disorders of aging, is a major contributor to vascular cognitive impairment and dementia (*6–8*). Neuroimaging markers of cSVD—including white matter hyperintensities, lacunes, and cerebral microbleeds—are highly prevalent in older adults and frequently occur in the absence of overt symptoms (*9*). Indeed, silent cerebrovascular lesions are several-fold more common than symptomatic stroke (*5*). Magnetic resonance imaging (MRI) is the primary modality used to detect these structural abnormalities, with fluid-attenuated inversion recovery (FLAIR) sequences identifying white matter hyperintensities and susceptibility-sensitive sequences revealing cerebral microbleeds (*9*). However, such structural markers often represent relatively late manifestations of vascular injury. Alterations in cerebral perfusion may precede visible structural changes, suggesting that quantitative assessment of CBF could provide an earlier indicator of cerebrovascular dysfunction (*10*).

Positron emission tomography (PET) using oxygen-15–labeled water ([^15^O]water) is considered the reference standard for quantitative measurement of regional CBF because of its freely diffusible tracer kinetics and well-established compartmental modeling (*11–14*). In clinical practice, however, [^15^O]water PET is used almost exclusively for quantitative assessment of myocardial blood flow in patients undergoing evaluation for suspected coronary artery disease (CAD) (*15*), whereas cerebral perfusion PET is rarely performed routinely as the main diagnostic imaging. The advent of long–axial field-of-view (laFOV) PET systems enables high-sensitivity dynamic imaging across multiple organs, such that the brain is inherently included in the FOV during e.g. cardiac [^15^O]water PET examinations and whole-body quantitative perfusion imaging can be obtained within a single scan (*16–18*). However, the clinical and research utility of such measurements depends on the availability of well-characterized reference ranges that account for normal physiological variability.

Indeed, despite this emerging opportunity, routine clinical interpretation of cerebral perfusion remains limited by the absence of well-characterized normative reference ranges for quantitative [^15^O]water PET across the adult lifespan and relevant physiological covariates. Although large-scale normative models of cerebral perfusion have recently been developed using arterial spin labeling (ASL) MRI (*19*), these reference values are not directly transferable to PET because of fundamental differences in acquisition and quantification methods. In PET, reported normal CBF values are typically derived from relatively small cohorts and heterogeneous imaging protocols, and measured CBF may vary depending on reconstruction and modeling approaches (*20*). Cerebral perfusion also varies with physiological factors such as age, sex, blood hemoglobin concentration, body composition, and metabolic state (*20–22*), emphasizing the need for covariate-adjusted reference frameworks.

The aim of the present study was to establish a normative, quantitative database of whole-brain cerebral blood flow across the adult lifespan using long-axial field-of-view PET and [^15^O]water. By incorporating key physiological covariates—including age, sex, hemoglobin concentration, and body mass index—we derived reference distributions using linear mixed effects models to describe expected variation in CBF. This framework provided quantitative reference curves for cerebral blood flow across the adult lifespan and supports automated identification of deviations from expected perfusion patterns.

## Material and Methods

### Study Subjects

The study combined data from four research projects conducted at Turku PET Centre between August 2022 and July 2025. All studies were performed using the same PET/CT scanner (Siemens Biograph Vision Quadra), imaging protocol, and image analysis pipeline. The pooled dataset consisted of participants undergoing quantitative cerebral perfusion imaging with [^15^O]water PET during resting or mild stimulation conditions. All studies complied with the Declaration of Helsinki. The Ethics Committee of the Hospital District of Southwest Finland approved the study protocols and written informed consent was provided by all study subjects. The largest subset consisted of patients referred for clinical myocardial perfusion imaging with [^15^O]water PET for evaluation of suspected coronary artery disease. Between August 2022 and July 2025, a total of 218 patients underwent rest and adenosine stress total-body PET imaging. Details of the adenosine stress protocol and multiorgan perfusion responses for a subset of the patients have been previously reported (*23*). For the present analysis, only the rest scans were included. In contrast with the previous analysis, which excluded patients based on cardiovascular findings, the current study excluded participants only if there was evidence of neurological disease. Individuals with previously diagnosed ischemic stroke, or other neurological disorders were excluded. Based on these criteria, 21 participants (47% female) were excluded. This study was registered in ClinicalTrials.gov (NCT05825859).

Additional data were obtained from three projects with volunteers recruited between October 2022 and October 2024. The first project investigated the effect of injected tracer dose on image quality and quantitative accuracy of [^15^O]water PET (ClinicalTrials.gov NCT06739473). Seven participants were scanned repeatedly at four dose levels within the same day. Six scans per participant were included in the present analysis after exclusion of the two lowest-dose scans for each participant due to insufficient count statistics. The second study investigated cerebral blood flow during emotional stimulation versus neutral baseline (ClinicalTrials.gov NCT06739473). Forty-four participants underwent repeated PET scans within the same day while either viewing neutral video stimuli or during mild cold exposure. In total, 172 scans (3–4 scans from the baseline condition per participant) were available. The third sample consisted of 53 healthy volunteers who each underwent one resting-state PET scan at room temperature and one under cold exposure. The CBF assessments at room temperature were included in the present study. In this study participants with high body mass index (BMI) were included in order to investigate the impact of adiposity to CBF (ClinicalTrials.gov NCT05468151). Across the three studies, all participants were adults without known neurological disease, as determined by participant self-report.

### PET Image Acquisition

All PET data were acquired using a long–axial field-of-view PET/CT scanner (Biograph Vision Quadra; Siemens Healthineers). Participants were instructed to abstain from caffeine for 24 hours prior to scanning. A low-dose CT scan was acquired for attenuation correction before PET imaging.

[^15^O]water was produced using an automated radiowater generator and administered as an intravenous bolus injection (target activity approximately 350 MBq, except for dose optimization study 100 – 700 MBq). Dynamic PET acquisition was initiated shortly after tracer administration and continued for four minutes and 40 seconds (cardiac patients) or seven minutes (volunteers). Images were reconstructed into dynamic frames (cardiac patients: 14 x 5 s, 3 x 10 s, 3 x 20 s, 4 x 30 s, and volunteers: 14 x 5 s, 3 x 10 s, 4 x 20 s, 4 x 30 s, and 2 x 60 s duration) using ultra–high-definition reconstruction in high-sensitivity mode with corrections for attenuation, scatter, randoms, and radioactive decay. Reconstruction parameters included 3 iterations and 5 subsets with a 3 mm Gaussian post-filter. These acquisition and reconstruction parameters were identical across all four contributing studies, ensuring methodological consistency of the quantitative perfusion measurements.

### PET Image Analysis

Image analysis was performed using a standardized automated processing pipeline developed at Turku PET Centre (TurBO), which has been previously validated for quantitative [^15^O]water PET perfusion imaging (*24*). Within the TurBO pipeline, brain preprocessing was conducted using the Magia toolbox, including motion correction (MAGIA) (*25*). CBF was quantified at each brain voxel using a one-tissue compartment model with an image-derived arterial input function extracted from the descending aorta (*24*). Perfusion values were expressed as milliliters of blood per minute per deciliter of tissue (mL/(min*dL)).

Region-wise perfusion values were extracted using the Desikan-Killiany brain atlas (*26*) after spatial normalization of the PET images. Spatial normalization was conducted using diffeomorphic *greedy* registration algorithm (*27*) and a high-resolution, in-house PET template as a target. Regional cortical and subcortical perfusion values were calculated for subsequent statistical analyses.

### Statistical Methods

Statistical analyses were performed using R (R version 4.4.3). Continuous variables are reported as mean ± SD unless otherwise stated. Group differences in demographic variables were assessed using unpaired t-tests. To evaluate the determinants of cerebral blood flow, linear mixed-effects (LME) models were fitted with age, sex, and body mass index (BMI) as fixed effects and subject as a random effect (random intercept) to account for repeated measurements within individuals (*28*). Age and BMI were mean-centered to facilitate interpretation of the model intercept. In a subset of participants with available hemoglobin (Hb) measurements, additional models were fitted including Hb as a covariate (n=221). Systolic and diastolic blood pressure (BP) were reported in a subset of participants (n=248).

To determine the appropriate functional form of covariate effects on CBF, models were fitted both on the original and log-scaled CBF values. The log-linear formulation assumes multiplicative effects corresponding to constant percentage changes in CBF, whereas the linear model assumes additive changes in absolute CBF units. Model performance was compared using residual mean squared error (RMSE), and the model exhibiting lower RMSE was selected for subsequent reporting of covariate effects.

For log-linear models, regression coefficients (β) were interpreted as percentage changes in CBF associated with a one-unit change in the covariate while holding the other covariates constant. Regional effects were evaluated across predefined brain regions of interest (*26*).

To enhance sensitivity to regional variation in cerebral blood flow (CBF), regional CBF values were in addition adjusted for individual differences in global grey matter CBF using a regression-based approach adapted from Buckner et al. (2004) (*31*). The rationale for this calibration was to enhance the detection of regional variations in CBF by reducing variability attributable to global perfusion differences. For each brain region, a linear regression model was fitted across subjects with regional CBF as the dependent variable and global grey matter CBF as the predictor. The resulting regression coefficient was then used to adjust individual regional CBF values by removing the component of regional perfusion attributable to deviations in global CBF from the sample mean. This procedure reduces global perfusion-related variability while preserving region-specific differences in CBF.

Multiple comparisons across brain regions were controlled using the Benjamini–Hochberg false discovery rate (FDR) procedure (*29*). Regional differences in effect sizes were evaluated by comparing 95% confidence intervals (95% CI). Variance components of the mixed-effects models were used to estimate between-subject and within-subject variability, expressed as coefficients of variation (CoV). Intraclass correlation coefficients (ICC) were calculated to assess the reliability of repeated CBF measurements.

Normative predictions of CBF across adulthood were generated using the fitted mixed-effects models. Prediction intervals (95%) were calculated to describe the expected range of CBF values conditional on age, sex, BMI, and hemoglobin. A two-sided P value < 0.05 was considered statistically significant.

### Data Availability

Voxelwise parameter maps of the normative models are available through NeuroVault. The generative model, example scripts, and visualization snapshots are available in an open repository (GitHub/Zenodo DOI), enabling prediction of expected cerebral blood flow and corresponding prediction intervals for individual subjects. The repository contains visualization snapshots and videos illustrating predicted cerebral perfusion across a range of physiological parameter values (age, sex, BMI, and hemoglobin). These resources enable researchers to reproduce the normative predictions and apply the generative models to independent datasets. The repository link will be provided upon publication. Per Finnish legislation, original imaging data are considered sensitive medical information and they cannot be distributed openly.

## Results

### Study demographics

Quantitative measures of cerebral blood flow using the same imaging parameters with [^15^O]water PET were obtained in 303 adults between ages 21 and 86 (N = 303, mean age 54.2±15.2 years). One participant’s data was excluded after outlier analysis (76-year-old female, spatial normalization failure despite multiple attempts).

Study demographics are presented in Table 1. The final sample included 139 females (n = 139, 46%, mean age 57.1±14.3 years) and 163 males (n = 163, 54%, mean age 51.7±15.5). BMI range was 19.5 kg/m^2^ to 55.5 kg/m^2^ (mean BMI 29.8±6.1 kg/m^2^), and was similar between sexes (p=0.39, unpaired t-test), and between younger vs. older participants (median split at 58.2 years, p=0.26, unpaired t-test). Of physiological parameters that may plausibly impact cerebral blood flow, hemoglobin (Hb) measurements were available in a subset of participants (n = 221, 55% female, mean age 58.5±13.5 years). In line with past findings (*30*), Hb was higher in men versus women (difference in mean Hb 15.4 g/L, p < 10^-17^, unpaired t-test), while no age effect was observed (p = 0.41, unpaired t-test). Furthermore, blood pressure was measured in a subset of participants (n = 248, 56% female, mean age 58.1±13.5 years). Systolic blood pressure range was 75.0 mmHg to 202 mmHg (mean systolic BP 140.4±22.1 mmHg). Blood pressure showed no difference between men and women, while there was significant elevation of systolic, but not diastolic blood pressure with advancing age (median split at 58.2 years, p<0.00001, unpaired t-test). However, despite systolic BP being significantly higher in the older age segment, it was only mildly elevated (mean systolic BP 145.8±22.8 mmHg) in this age group.

**Table 1:** Demographics of the study (mean±SD).

|  | All | Female | Male | <58.2 | >58.2 |
| --- | --- | --- | --- | --- | --- |
| Participants | 302 | 139 | 163 | 151 | 151 |
| Mean age | 54.2±15.1 | 57.1±14.3 | 51.7±15.5 | 41.4±9.8 | 67.0±5.8 |
| Age range | 21.0-85.7 | 22.0-81.1 | 21.0-85.7 | 21.0-58.1 | 58.2-85.7 |
| Mean BMI (kg/m<sup>2</sup>) | 29.8±6.1 | 29.5±6.1 | 30.1±6.1 | 29.4±7.0 | 30.2±5.0 |
| BMI range (kg/m<sup>2</sup>) | 19.5-55.6 | 19.5-52.6 | 20.1-55.6 | 19.5-55.6 | 21.0-48.4 |
| Mean Hb (g/L) | (n=221)<br>141.4±13.8 | (n=121)<br>134.4±10.8 | (n=100)<br>149.8±12.5 | (n=86)<br>139.3±15.4 | (n=135)<br>142.8±12.7 |
| Hb range (g/L) | 82.0-174.0 | 82.0-161.0 | 99.0-174.0 | 82.0-171.0 | 100.0-174.0 |
| Mean systolic BP (mmHg) | (n=248)<br>140.4±22.1 | (n=139)<br>141.4±22.8 | (n=109)<br>139.2±21.1 | (n=97)<br>132.1±18.1 | (n=151)<br>145.8±22.8 |
| Systolic BP range (mmHg) | 75.0-202.0 | 96.0-201.0 | 75.0-202.0 | 103.5-191.0 | 75.0-202.0 |
| Mean diastolic BP (mmHg) | (n=248)<br>81.1±12.2 | (n=139)<br>81.9±12.2 | (n=109)<br>80.3±12.2 | (n=97)<br>81.1±11.7 | (n=151)<br>81.2±12.5 |
| Diastolic BP range (mmHg) | 42.0-130.0 | 55.0-130.0 | 42.0-108.0 | 53.0-111.0 | 42.0-130.0 |

### Average cerebral blood flow in the grey matter is approximately 50 mL/min*dL

Linear mixed effects models with age, sex and BMI as fixed effects and subject as random effect (random intercept) were used to predict CBF. Age and BMI were mean-centered so that the intercept represents the expected CBF for a (female) participant with average age and BMI in the sample (average age 54.2 years, BMI 29.8 kg/m^2^). Whole brain maps of model-derived average CBF are shown in Figure 1A, and regional summaries in Figure 1B and Supplemental Table 1. Average CBF was 46.1 mL/min*dL across all grey matter voxels in the brain. Regionally, average CBF was within a range of 29.2 mL/min*dL (temporal pole) to 58.2 mL/min*dL (putamen), translating to a 100% difference between the highest and lowest regional rates of CBF. The 95% prediction intervals (95%PI, Fig 1B, Supplemental Table 1) indicated large within-region variance in CBF across people. In average, the prediction intervals suggested a -26.8 % to 36.4 % predicted variance around the mean CBF (Fig 1B).

**Figure 1.**
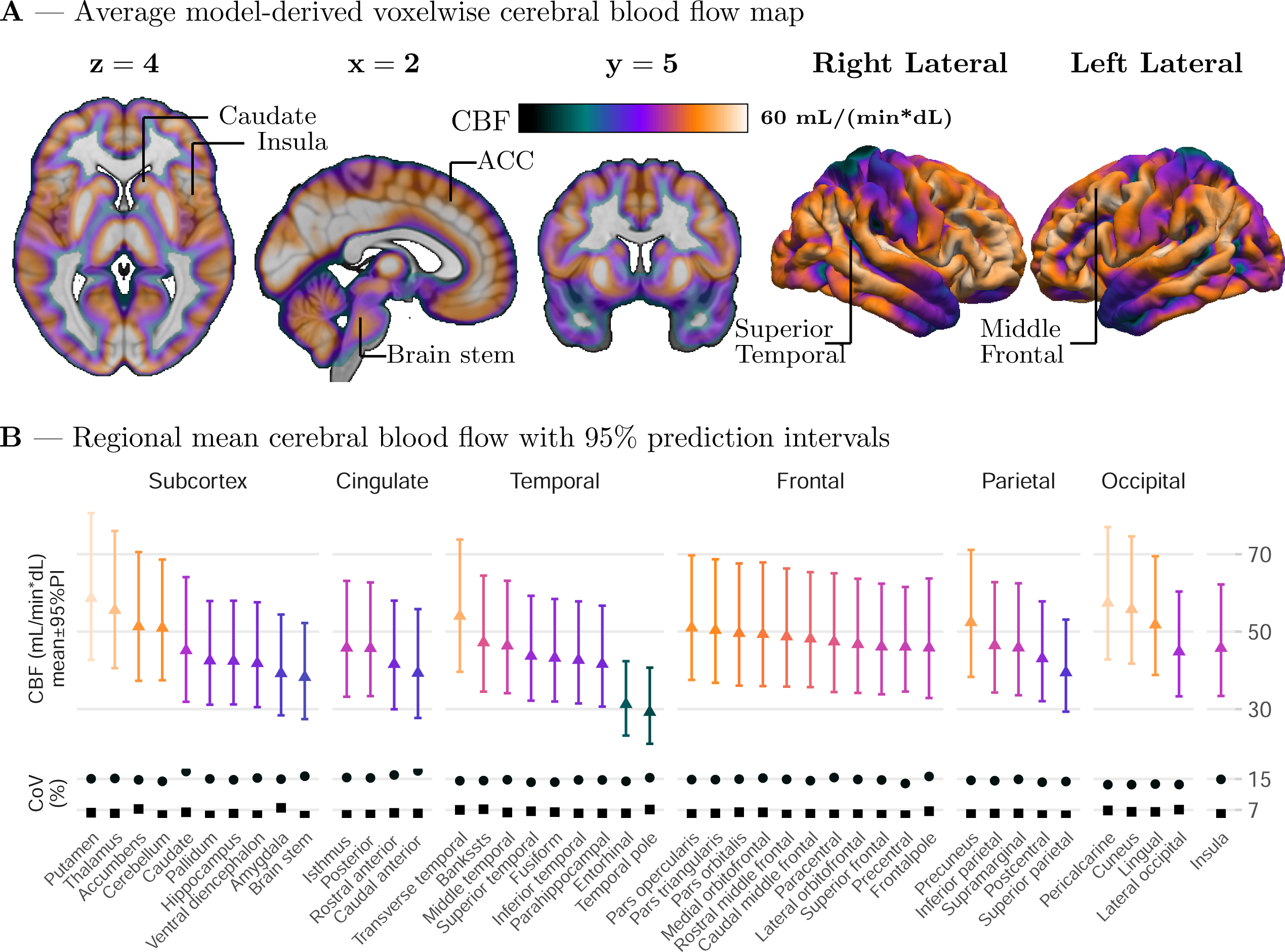
Average model-derived cerebral blood flow. (A) Whole-brain voxel-wise CBF map. (B) Regional mean CBF with 95% prediction intervals and coefficients of variation (CoV%) reflecting between- (circles) and within-subject (squares) variability. ACC, anterior cingulate cortex.

We next decomposed CBF variance in terms of between- and within-person variances. Past research has indicated large variation across subjects (*19*,*20*), yet within-person variances have typically not been reported in the same study. Hence, it is unclear how much the large CBF variance is due to methodological versus true biological variation. Repeated measures of CBF were available in a subset of participants (n=51, 73% female, age range 21 – 62 years, n_scans_=214), permitting the calculation of LME-derived estimates of between- versus within-person variances. The results indicated small regional differences in between-subject variances (Fig 1B, Supplemental Table 1, Supplemental Fig 1), with all regional between-subject coefficients of variation (CoV%) falling within a range of 13.5 % to 17.1 % (Fig 1B, Supplemental Table 1). In turn, within-subject CoV% were within a range of 5.5 % to 7.5 % (CoV%) indicating good methodological replicability (stability) of radio-water derived estimates of CBF. In keeping with these results, model-derived intra-class correlation coefficients (ICC) were high and consistent across all regions (ICC range 0.78 – 0.89, Supplemental Table 1, Supplemental Fig 1).

### Effects of age, sex, and BMI on cerebral blood flow

We next tested for the effects of age, sex and BMI on CBF. We first conducted a model comparison between linear mixed-effects models fitted for the original versus log-transformed data to evaluate two alternative forms of predictor effects on CBF. A log-linear model implies multiplicative effects, whereas a linear model assumes additive effects on CBF. Both models explained variance in the data (mean marginal R^2^ = 0.39, range 0.26 – 0.51). Model comparison indicated overall superior residual mean squared error (RMSE) for log-scaled LMEs (mean RMSE difference 0.012, RMSE_log_<RMSE_linear_ in 67% or regions). However, in the caudate and occipital and cingulate cortices the RMSE comparison favored a linear model (mean RMSE differences -0.010, -0.038, and -0.013, respectively), indicating that covariate-related differences increased in relative (percentage) terms across the observed range. Despite this marginal model fit disadvantage in some regions, the log-linear model was selected as the representative model for reporting the effects of the covariates. Whole brain maps of age-, sex- and BMI-related effects to CBF are presented in Fig 2, and regional summaries are presented in Fig 2 and Supplemental Table 2. In the log-linear model, a one-unit change in a covariate corresponds to a percentage change in CBF. Accordingly, the regression coefficients (β) were interpreted as the percentage change in CBF associated with a one-unit change in the covariate, holding the other covariates constant.

**Figure 2.**
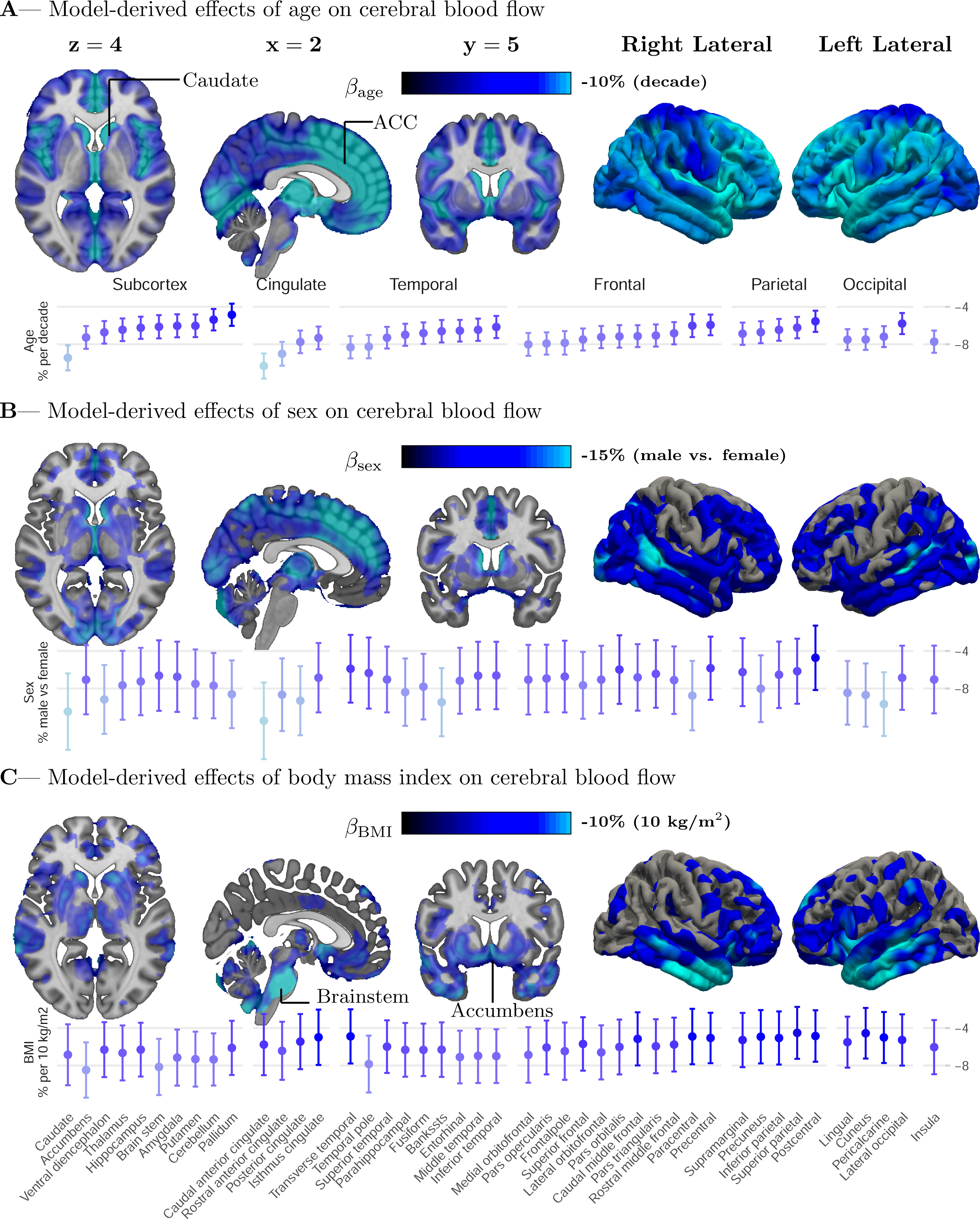
Model-derived estimates of age-, sex-, and BMI-related differences in cerebral blood flow (A–C). Voxel-wise β maps with corresponding regional summaries for age (A), sex (B), and BMI (C). Error bars display 95% confidence intervals. ACC, anterior cingulate cortex.

Regional analysis indicated a negative effect of advancing age to CBF in all brain regions (p_FDR_ < 10^-12^, Fig 2A, Supplemental Table 2). Whole brain maps suggest regional differences in the effects of age, with more pronounced effects in the midline regions (Fig 2A). Mean rate of CBF decline was approximately 7% per decade (0.7% per year; Supplemental Table 2), while more prominent decline of 10% per decade was observed in the caudate and anterior cingulate cortex (Fig 2A, Supplemental Table 2). Statistically significant regional differences were defined using non-overlapping 95% confidence intervals (95%CI, Fig 2A, Supplemental Table 2). This criterion was met in the caudate in comparison to 50% of all brain regions, in 80% of subcortical regions, and the entire parietal cortex (Fig 2A, Supplemental Table 2). In terms of caudal anterior cingulate cortex (ACC), CBF reductions were significantly larger in comparison to 82% of brain regions (Fig 2A, Supplemental Table 2).

### Sex-related CBF differences and hemoglobin

Regional analysis indicated that females had higher CBF than males in all brain regions (p_FDR_ < 0.01, Fig 2B, Supplemental Table 2). Whole brain maps suggest potential regional differences in the effects of sex (Fig 2B), which were however not statistically significant using the non-overlapping 95%CI criteria for regional data (Fig 2B, Supplemental Table 2). In average, men exhibited approximately 7.5% [95%CI -11.1% - -3.9%] lower CBF relative to women consistently across the entire brain (Fig 2B, Supplemental Table 2). Follow-up analyses indicated no interaction between age and sex (p_FDR_ > 0.5), suggesting a stable negative association between male sex and CBF across adulthood.

We next investigated the role of sex-related hemoglobin differences in CBF, as lower blood hemoglobin in women might be associated with elevated CBF to yield similar oxygen delivery to brain tissue across sexes (*22*). To that end, the subset of participants with hemoglobin measurements was included in a follow-up analyses (c.f. Table 1). We observed statistically significantly higher Hb in men versus women (n=221, difference in mean Hb 15.4 g/L, p < 10^-17^, unpaired t-test, Table 1). Furthermore, in the subset, men exhibited approximately 10% [95%CI - 14.8% - -6.0%] lower CBF relative to women across the entire brain (p_FDR_ < 0.001). Inclusion of Hb in the LME model almost entirely abolished sex-related differences (p_FDR_ > 0.05 in 95% of regions), with still significantly higher CBF in females confined to the posterior and caudal anterior cingulate cortices (p_FDR_ = 0.033). Age-related CBF differences were not affected by inclusion of Hb in the model (p_FDR_ < 0.001).

### Effects of BMI on CBF

Regional analysis indicated a negative effect of BMI on CBF in all brain regions (p_FDR_ < 0.01, Fig 2C, Supplemental Table 2). Whole brain maps suggest potential regional differences in the effects of BMI (Fig 2C), which were however not statistically significant using the non-overlapping 95%CI criteria (Fig 2C, Supplemental Table 2). On average, higher BMI was associated with approximately 6% [95%CI -9.0% - -3.2%] lower CBF per 10kg/m^2^ (Fig 2C, Supplemental Table 2). Largest effects of approximately 8% per 10kg/m^2^ were observed in the accumbens area and brain stem, even though the 95%CI were overlapping with other brain regions (Fig 2C, Supplemental Table 2). Follow-up analyses indicated no interaction between age and BMI (ps > 0.3), suggesting a stable negative association between heightened BMI and CBF across adulthood. Furthermore, inclusion of Hb in the LME model did not significantly influence the effect of BMI to CBF (ps < 0.05).

### Normative whole-brain CBF maps across the adult life span

We finally trained generative models of whole-brain CBF across the adult life span. Two models were generated to accommodate 1) age-, sex-, and BMI-related differences and 2) additionally hemoglobin-related differences. The primary model excluded hemoglobin as a covariate due to the frequent omission of hemoglobin assessment in CBF studies; the rationale was to offer models with minimal set of parameters to mitigate the impact of missing data. Figure 3A shows whole-brain voxel-wise maps of predicted CBF across a range of age, BMI and hemoglobin and between sexes. Prediction intervals as a function of age are shown in Figure 3B. The predicted CBF across realistic range of age, BMI and Hb illustrate the impact of change in each variable relative to the other variables (Fig 3A). According to Fig 3B the width of 95% prediction intervals remained relatively constant across the adult lifespan. However, the negative age-related differences in CBF were so dramatic that over half of the elderly participants (>65 y) exhibited CBF rates below 95%PI of a young person (age 20 y; Fig 3B). Hence, age-matching is imperative in the assessment of an individuals CBF relative to the normative CBF maps. Similarly, individuals’ BMI and Hb have impact on the expected CBF rate, influencing the interpretation. These effects are, however, in general modest.

**Figure 3.**
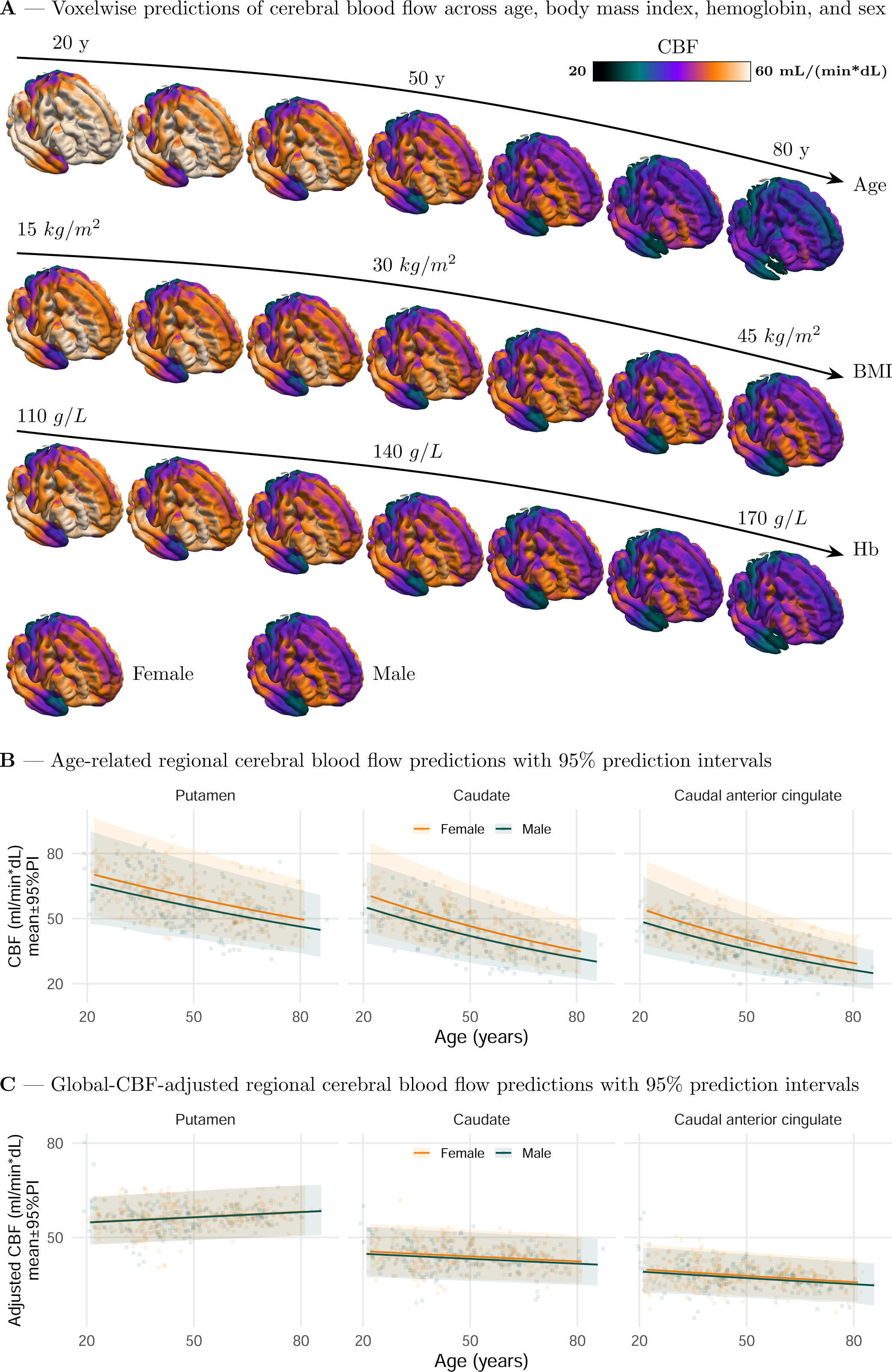
Voxel-wise and regional predictions of cerebral blood flow across demographic, anthropometric, and physiological factors. (A) Whole-brain voxel-wise maps showing model-predicted cerebral blood flow (CBF) across the adult lifespan, varying levels of adiposity, and hemoglobin concentration. (B) Corresponding regional predictions of CBF summarized across predefined brain regions, displayed as mean estimates with 95% prediction intervals. (C) Global-CBF–adjusted regional predictions, demonstrating substantially narrower 95% prediction intervals compared with unadjusted regional estimates.

Finally, we adjusted individuals’ CBF by their global grey matter CBF (*31*), and recalculated the model parameters. The rationale for calibrating by global CBF was to enhance the detection of local variances in CBF. Results for global-CBF-adjusted analysis in selected regions are shown in Fig 3C. Expectedly, the prediction intervals were narrower after adjustment for global CBF (Fig 3C). The mean width of prediction intervals were 29.5 mL/min*dL and 12.7 mL/min*dL for original and adjusted CBFs, respectively. Together, the present results suggest that global-CBF-adjustment may improve the sensitivity to detect regional variations in CBF due to e.g. stroke.

## Discussion

In this study we established a normative database of quantitative cerebral blood flow (CBF) across the adult lifespan using total-body [^15^O]water PET in a cohort of over 300 neurologically healthy adults. Average grey matter CBF was approximately 46 mL/min*dL, with substantial regional and inter-individual variability, but good within-subject replicability. Advancing age and higher body mass index were consistently associated with lower CBF across brain regions. Women exhibited higher CBF than men, but most sex-related differences were explained by differences in hemoglobin concentration, likely related to compensatory changes in perfusion that help maintain oxygen delivery to brain tissue. Using these data, we also generated covariate-adjusted normative models describing expected CBF across adulthood as a function of age, sex, BMI, and hemoglobin. These normative models provide a quantitative reference framework for interpreting cerebral perfusion and may facilitate automated detection of abnormal CBF in patients. This is particularly important given the advent of long axial field of view PET imaging, where brain data can be acquired alongside with the main target organ (e.g. heart), thus assisting in detection of incidental cerebral findings (c.f. Knuuti et al. 2023).

### Demographic factors influencing cerebral perfusion

Our estimates of absolute cerebral blood flow are broadly consistent with previous measurements obtained using PET (*20*) and ASL MRI (*19*). Across all grey matter voxels, mean CBF was approximately 46 mL/min*dL, which falls within the range reported in classical studies of healthy adults (*32*). Regional variation was also consistent with known perfusion patterns, with higher CBF observed in subcortical nuclei and lower values in anterior temporal regions (*19*). Notably, substantial inter-individual variability in CBF was observed across participants, with prediction intervals spanning approximately ±30% around the regional mean, also in accord with past findings (*19*,*33*,*34*). Decomposition of variance indicated that most of this variability reflected between-subject differences rather than measurement-related variability, as within-subject coefficients of variation were small and intraclass correlation coefficients were high across regions, in line with past findings (*35*). Together, these findings support the robustness of quantitative total-body [^15^O]water PET for measuring cerebral perfusion and highlight the importance of accounting for biological variability when interpreting individual CBF measurements.

Advancing age was associated with a consistent decline in cerebral blood flow across the entire brain, with an average reduction of approximately 7% per decade. This estimate is in close agreement with previous PET studies reporting progressive reductions in CBF with aging, generally on the order of 5–10% per decade in healthy adults (*20*,*33*). Although the age-related decline was observed in all regions, modest regional differences were detected, with the largest effects in the caudate and anterior cingulate cortex. Such patterns are consistent with previous observations that subcortical and midline structures may be particularly sensitive to age-related vascular and molecular changes (*20*,*36*). Importantly, the magnitude of the age effect was sufficiently large that predicted CBF values for many older individuals fell below the normative range of younger adults, emphasizing the importance of age-matched reference values when interpreting quantitative perfusion measurements. These findings highlight aging as a major determinant of inter-individual variability in CBF and underscore the need for lifespan-adjusted normative models when evaluating cerebral perfusion in clinical populations.

Females exhibited higher CBF than males across the brain; however, this difference was largely explained by hemoglobin concentration. Hemoglobin is the principal determinant of arterial oxygen content and an important regulator of cerebral perfusion. PET studies have demonstrated an inverse relationship between hemoglobin (or hematocrit) and CBF, reflecting a compensatory adjustment of perfusion to maintain stable oxygen delivery to brain tissue (*34*,*37*). In particular, Ibaraki and colleagues showed using ^15^O PET that CBF and oxygen extraction fraction vary systematically with hemoglobin concentration, whereas the cerebral metabolic rate of oxygen remains relatively stable across individuals (*34*), consistent with homeostatic regulation of cerebral oxygen metabolism. The present results align with this physiological framework: women exhibited higher raw CBF than men, but the difference was largely abolished after accounting for hemoglobin. Commonly reported sex differences in cerebral perfusion may thus largely reflect hematologic differences in oxygen-carrying capacity rather than intrinsic differences in cerebrovascular regulation. This observation highlights hemoglobin as an important covariate when interpreting quantitative CBF measurements, particularly in studies comparing perfusion across populations with systematic differences in blood oxygen-carrying capacity.

Higher BMI was associated with lower CBF across the brain, with an average reduction of approximately 6% per 10 kg/m². Only a few prior PET studies have examined this relationship, but our findings are broadly consistent with MRI-based perfusion studies reporting lower cerebral perfusion in individuals with higher BMI or obesity (*38*,*39*). Several mechanisms may contribute to this association. Obesity is linked to vascular and metabolic alterations—including endothelial dysfunction, chronic inflammation, insulin resistance, and increased arterial stiffness—that may impair cerebrovascular regulation and reduce cerebral perfusion (*40*). Notably, the BMI effect observed in the present study was relatively uniform across brain regions and independent of age and hemoglobin, suggesting that increased body mass exerts a global influence on cerebral perfusion rather than a strongly region-specific effect. These results further highlight BMI as an important physiological covariate to consider when interpreting quantitative CBF measurements in both research and clinical settings.

### Normative models of CBF

A central aim of the present study was to establish covariate-adjusted normative models for quantitative cerebral blood flow across the adult lifespan. Our results demonstrate that age, sex, BMI, and hemoglobin account for a substantial portion of inter-individual variability in CBF, highlighting the importance of incorporating physiological covariates when interpreting perfusion measurements. The resulting models provide predicted CBF values and corresponding prediction intervals across adulthood, enabling individual measurements to be evaluated relative to expected values for a person with comparable physiological characteristics. In particular, the pronounced age-related decline in CBF observed here is consistent with prior PET studies of aging (*20*,*33*) and emphasizes that age-matched reference values are essential for accurate interpretation of perfusion measurements. These normative frameworks are particularly relevant in the context of long–axial field-of-view PET imaging, which enables high-sensitivity dynamic imaging across the entire body (*16*,*17*). Because the brain is inherently included during cardiac [^15^O]water PET examinations, cerebral perfusion data can be obtained without additional scanning or radiation exposure. The normative models presented here provide a quantitative reference framework that may facilitate automated detection of abnormal cerebral perfusion in patients undergoing PET imaging for suspected coronary artery disease, potentially enabling earlier identification of silent cerebrovascular pathology. Although [^15^O]water PET is considered the reference standard for quantitative perfusion measurement (*11*), methodological differences across scanners, reconstruction methods, or kinetic modeling approaches may influence absolute CBF estimates and should be considered when applying these normative values to other datasets.

## Limitations

The study population included adults drawn from cardiac PET studies rather than from a population-based cohort, which may limit generalizability to the broader population. Hemoglobin measurements were available only for a subset of participants, which reduced statistical power for analyses involving this covariate and may have limited detection of residual sex-related differences in perfusion. Although repeated scans were available in a subset of participants enabling estimation of within-subject variability, most individuals contributed a single measurement, resulting in incomplete coverage of within-subject variability across the full sample. Additional physiological factors that may influence cerebral perfusion—such as blood pressure, arterial CO_2_ levels, or medication use—were not systematically available for all participants.

## Conclusions

The present study establishes a normative database of quantitative cerebral blood flow across the adult lifespan using total-body [^15^O]water PET. Cerebral perfusion exhibited substantial inter-individual variability but showed consistent associations with age, BMI, and hemoglobin concentration. Incorporating these covariates into normative models enabled generation of quantitative reference curves describing expected CBF across adulthood. Given that long–axial field-of-view PET enables simultaneous imaging of the brain and heart during routine PET examinations, these models provide a framework for interpreting cerebral perfusion measurements and may support automated identification of abnormal CBF in clinical populations at risk.

## Data Availability

All data produced in the present study are available upon reasonable request to the authors

## Acknowledgements

This work was supported by European Research Council (ERC-ADV #101141656), Jane and Aatos Erkko Foundation, Gyllenberg’s Stiftelse, and The Finnish Governmental Research Funding for Turku University Hospital, Finnish Cultural Foundation, Finnish Foundation for Cardiovascular Research and for the Western Finland collaborative area, Finnish Diabetes Research Foundation, Sigrid Juselius Foundation, Academy of Finland (343410), and the Research Council of Finland.

Dr Johansson has been supported/funded by the European Union’s Horizon Europe Framework programme for research and innovation 2021-2027 under the Marie Skłodowska-Curie grant agreement No 101126611.

---

**Supplemental Table 1:** Representative regional estimates of CBF for a female participant with average age and BMI in the sample (age 54.2 years, BMI 29.8 kg/m^2^), and model-derived estimates of regional coefficients of variance (CoV) and intra-class correlation coefficients (ICC). Despite a large regional variation in CBF, the model-derived variance parameters indicated fairly homogeneous intra- and inter-individual variations of CBF across regions.

| **Lobe** | **Region** | **CBF (mL/(min\*dL))**<br>**[95% PI]** | **CoV (%)**<br>**between** | **CoV (%)**<br>**within** | **ICC** |
| --- | --- | --- | --- | --- | --- |
| Subcortex | Putamen | 58.18 [42.80-81.42] | 15.07 | 6.29 | 0.85 |
| Subcortex | Thalamus | 55.00 [40.62-75.66] | 15.15 | 6.08 | 0.86 |
| Subcortex | Cerebellum | 50.80 [37.10-67.80] | 14.39 | 5.67 | 0.86 |
| Subcortex | Accumbens | 50.73 [37.18-71.03] | 14.78 | 7.25 | 0.80 |
| Subcortex | Caudate | 45.04 [31.40-63.43] | 16.93 | 6.42 | 0.87 |
| Subcortex | Pallidum | 42.26 [30.97-58.01] | 15.03 | 5.59 | 0.88 |
| Subcortex | Hippocampus | 42.24 [30.30-57.81] | 14.78 | 6.05 | 0.86 |
| Subcortex | Ventral diencephalon | 41.73 [30.72-56.92] | 15.26 | 5.66 | 0.88 |
| Subcortex | Amygdala | 39.37 [28.46-53.58] | 14.95 | 7.55 | 0.80 |
| Subcortex | Brain stem | 38.43 [26.21-52.25] | 15.72 | 5.52 | 0.89 |
| Frontal | Pars opercularis | 51.16 [37.62-68.59] | 14.84 | 5.97 | 0.86 |
| Frontal | Pars triangularis | 50.18 [36.29-68.03] | 14.83 | 6.10 | 0.85 |
| Frontal | Pars orbitalis | 49.91 [36.79-66.74] | 14.96 | 6.43 | 0.84 |
| Frontal | Medial orbitofrontal | 49.14 [36.27-67.12] | 15.24 | 6.39 | 0.85 |
| Frontal | Rostral middle frontal | 48.22 [35.38-64.96] | 14.90 | 5.82 | 0.87 |
| Frontal | Caudal middle frontal | 47.91 [35.46-65.25] | 14.56 | 6.02 | 0.85 |
| Frontal | Paracentral | 47.18 [34.50-63.96] | 15.37 | 5.85 | 0.87 |
| Frontal | Lateral orbitofrontal | 46.31 [34.69-64.03] | 14.88 | 5.92 | 0.86 |
| Frontal | Precentral | 45.96 [34.20-61.99] | 13.84 | 5.66 | 0.86 |
| Frontal | Superior frontal | 45.70 [33.77-63.44] | 14.74 | 5.75 | 0.87 |
| Frontal | Frontalpole | 45.18 [33.11-62.24] | 15.65 | 6.69 | 0.84 |
| Parietal | Precuneus | 52.41 [38.01-70.70] | 14.65 | 5.93 | 0.86 |
| Parietal | Inferior parietal | 46.73 [33.66-62.96] | 14.55 | 6.07 | 0.85 |
| Parietal | Supramarginal | 46.05 [33.95-61.56] | 14.91 | 6.09 | 0.86 |
| Parietal | Postcentral | 42.81 [31.62-58.35] | 14.16 | 5.69 | 0.86 |
| Parietal | Superior parietal | 39.50 [29.43-52.59] | 14.34 | 5.60 | 0.87 |
| Temporal | Transverse temporal | 53.53 [39.75-71.99] | 14.54 | 7.01 | 0.81 |
| Temporal | Bankssts | 46.98 [34.27-64.11] | 14.57 | 7.19 | 0.80 |
| Temporal | Middle temporal | 46.10 [33.74-62.14] | 14.80 | 6.36 | 0.84 |
| Temporal | Superior temporal | 43.79 [31.85-58.60] | 14.16 | 6.67 | 0.82 |
| Temporal | Fusiform | 43.09 [32.26-58.55] | 14.19 | 6.45 | 0.83 |
| Temporal | Inferior temporal | 42.40 [31.19-58.73] | 14.76 | 6.12 | 0.85 |
| Temporal | Parahippocampal | 41.66 [30.83-56.65] | 14.75 | 6.12 | 0.85 |
| Temporal | Entorhinal | 31.19 [23.23-42.23] | 14.40 | 6.15 | 0.84 |
| Temporal | Temporal pole | 29.19 [20.80-40.39] | 15.33 | 7.13 | 0.82 |
| Occipital | Pericalcarine | 56.99 [42.54-76.40] | 13.54 | 6.89 | 0.79 |
| Occipital | Cuneus | 55.82 [41.48-74.32] | 13.55 | 6.52 | 0.81 |
| Occipital | Lingual | 51.60 [38.59-69.31] | 13.69 | 6.51 | 0.81 |
| Occipital | Lateral occipital | 44.45 [33.44-60.66] | 13.57 | 7.09 | 0.78 |
| Cingulate | Posterior cingulate | 45.51 [33.79-63.48] | 15.27 | 5.81 | 0.87 |
| Cingulate | Isthmus cingulate | 45.33 [34.16-62.43] | 15.38 | 5.73 | 0.88 |
| Cingulate | Rostral anterior cingulate | 41.26 [29.87-57.49] | 16.04 | 6.25 | 0.87 |
| Cingulate | Caudal anterior cingulate | 38.80 [27.35-55.24] | 17.08 | 6.10 | 0.89 |
| Insula | Insula | 46.06 [33.58-63.40] | 14.90 | 5.97 | 0.86 |

**Supplemental Table 2:**
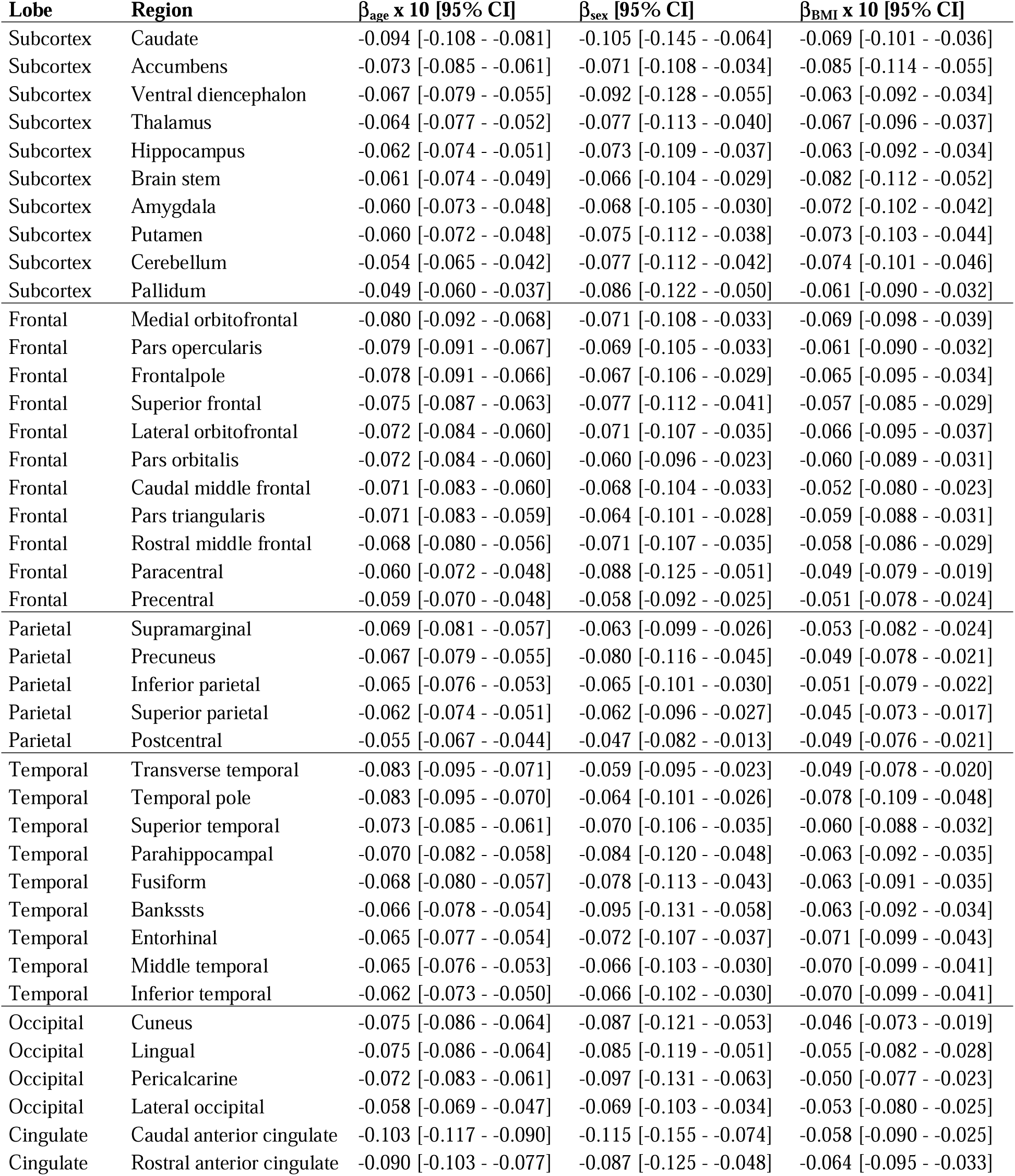
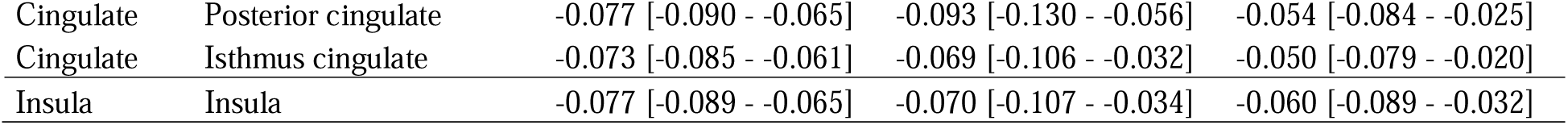
Effects of age, sex (reference level is female; negative beta means male < female), and BMI on CBF. Model coefficients (betas) correspond to percentage change in CBF per unit change in the covariate. Coefficients for age and BMI were multiplied by 10 to obtain percentage **change** in CBF in a decade or 10 kg/m^2^, respectively. All model terms were statistically significant (p_FDR_ < 0.01).

## References

1. Raichle ME, Gusnard DA. Appraising the brain’s energy budget. Proceedings of the National Academy of Sciences. 2002;99:10237–10239.

2. Willie CK, Tzeng YC, Fisher JA, Ainslie PN. Integrative regulation of human brain blood flow. Journal of Physiology. 2014;592:841–859.

3. Dirnagl U, Iadecola C, Moskowitz MA. Pathobiology of ischaemic stroke: an integrated view. Trends Neurosci. 1999;22.

4. Feigin VL, Stark BA, Johnson CO, et al. Global, regional, and national burden of stroke and its risk factors, 1990–2019: a systematic analysis for the Global Burden of Disease Study 2019. Lancet Neurol. 2021;20:795–820.

5. Vermeer SE, Longstreth WT, Koudstaal PJ. Silent brain infarcts: a systematic review.; 2007.

6. Pantoni L. Cerebral small vessel disease: from pathogenesis and clinical characteristics to therapeutic challenges. Lancet Neurol. 2010;9:689–701.

7. Iadecola C. The Pathobiology of Vascular Dementia. Neuron. 2013;80:844–866.

8. Vermeer SE, Prins ND, den Heijer T, et al. Silent Brain Infarcts and the Risk of Dementia and Cognitive Decline. Vol 13.; 2003.

9. Wardlaw JM, Smith EE, Biessels GJ, et al. Neuroimaging standards for research into small vessel disease and its contribution to ageing and neurodegeneration. Vol 12.; 2013.

10. Smith EE, Saposnik G, Biessels GJ, et al. Prevention of Stroke in Patients with Silent Cerebrovascular Disease: A Scientific Statement for Healthcare Professionals from the American Heart Association/American Stroke Association. Stroke. 2017;48:e44–e71.

11. Herscovitch P, Markham J, Raichle ME. Brain blood flow measured with intravenous H2(15)O. I. Theory and error analysis. Journal ofNuclear Medicine. 1983;9:782–789.

12. Åhlström A, Lindström E, Maaniitty T, et al. Automated Total-Body Perfusion Imaging with 15O-Water PET Using Basis Functions and Organ-Specific Model Selection. J Nucl Med. 2025;66:1307–1313.

13. Raichle ME, Martin WR, Herscovitch P, Mintun MA, Markham J. Brain blood flow measured with intravenous H2(15)O. II. Implementation and validation. J Nucl Med. 1983;24:790–8.

14. Kanno I, Iida H, Miura S, Murakami M. Optimal scan time of oxygen-15-labeled water injection method for measurement of cerebral blood flow. J Nucl Med. 1991;32:1931–4.

15. Kajander SA, Joutsiniemi E, Saraste M, et al. Clinical value of absolute quantification of myocardial perfusion with 15O-water in coronary artery disease. Circ Cardiovasc Imaging. 2011;4:678–684.

16. Cherry SR, Jones T, Karp JS, Qi J, Moses WW, Badawi RD. Total-body PET: Maximizing sensitivity to create new opportunities for clinical research and patient care. Journal of Nuclear Medicine. 2018;59:3–12.

17. Badawi RD, Shi H, Hu P, et al. First human imaging studies with the explorer total-body PET scanner. Journal of Nuclear Medicine. 2019;60:299–303.

18. Knuuti J, Tuisku J, Kärpijoki H, et al. Quantitative Perfusion Imaging with Total-Body PET. Journal of Nuclear Medicine. 2023;64:11S–19S.

19. Farahani A, Liu ZQ, Ceballos EG, et al. Mapping cerebral blood perfusion and its links to multi-scale brain organization across the human lifespan. PLoS Biol. 2025;23.

20. Leenders KL, Perani D, Lammertsma AA, et al. Cerebral Blood Flow, Blood Volume and Oxygen Utilization: Normal Values and Effect of Age. Brain. 1990;113:27–47.

21. Gur RE, Gur RC. Gender Differences in Regional Cerebral Blood Flow. Vol 16.; 1990.

22. van der Veen PH, Muller M, Vincken KL, et al. Hemoglobin, hematocrit, and changes in cerebral blood flow: The Second Manifestations of ARTerial disease-Magnetic Resonance study. Neurobiol Aging. 2015;36:1417–1423.

23. Kärpijoki H, Tuisku J, Palonen S, et al. Organ-Specific Perfusion Response to Adenosine as Measured Using Total-Body PET. Journal of Nuclear Medicine. February 2026:jnumed.125.271613.

24. Tuisku J, Palonen S, Kärpijoki H, et al. Automated long axial field of view PET image processing and kinetic modelling with the TurBO toolbox. Eur J Nucl Med Mol Imaging. January 2026.

25. Karjalainen T, Tuisku J, Santavirta S, et al. Magia: Robust Automated Image Processing and Kinetic Modeling Toolbox for PET Neuroinformatics. Front Neuroinform. 2020;14.

26. Desikan RS, Ségonne F, Fischl B, et al. An automated labeling system for subdividing the human cerebral cortex on MRI scans into gyral based regions of interest. Neuroimage. 2006;31:968–980.

27. Venet L, Pati S, Feldman MD, Nasrallah MP, Yushkevich P, Bakas S. Accurate and robust alignment of differently stained histologic images based on greedy diffeomorphic registration. Applied Sciences (Switzerland). 2021;11:1–18.

28. Bates D, Mächler M, Bolker BM, Walker SC. Fitting linear mixed-effects models using lme4. J Stat Softw. 2015;67.

29. Benjaminit Y, Hochberg Y. Controlling the False Discovery Rate: a Practical and Powerful Approach to Multiple Testing. Vol 57.; 1995.

30. Beutler E, Waalen J. The definition of anemia: what is the lower limit of normal of the blood hemoglobin concentration? 2006.

31. Buckner RL, Head D, Parker J, et al. A unified approach for morphometric and functional data analysis in young, old, and demented adults using automated atlas-based head size normalization: Reliability and validation against manual measurement of total intracranial volume. Neuroimage. 2004;23:724–738.

32. Lassen NA. Normal Average Value of Cerebral Blood Flow in Younger Adults is 50 ml/100 g/min. Journal of Cerebral Blood Flow & Metabolism. 1985;5:347–349.

33. Ito H, Kanno I, Kato C, et al. Database of normal human cerebral blood flow, cerebral blood volume, cerebral oxygen extraction fraction and cerebral metabolic rate of oxygen measured by positron emission tomography with 15O-labelled carbon dioxide or water, carbon monoxide and oxygen: A multicentre study in Japan. Eur J Nucl Med Mol Imaging. 2004;31:635–643.

34. Ibaraki M, Shinohara Y, Nakamura K, Miura S, Kinoshita F, Kinoshita T. Interindividual variations of cerebral blood flow, oxygen delivery, and metabolism in relation to hemoglobin concentration measured by positron emission tomography in humans. Journal of Cerebral Blood Flow and Metabolism. 2010;30:1296–1305.

35. Bremmer JP, Van Berckel BNM, Persoon S, et al. Day-to-day test-retest variability of CBF, CMRO2, and OEF measurements using dynamic 15O PET studies. Mol Imaging Biol. 2011;13:759–768.

36. Johansson J, Karalija N, Salami A. Cerebrovascular integrity affects gradients of aging-related dopamine D1 differences in the striatum. Aging Brain. 2023;4.

37. Kusunoki M, Kimura K, Nakamura M, Isaka Y, Yoneda S, Abe H. Effects of hematocrit variations on cerebral blood flow and oxygen transport in ischemic cerebrovascular disease. J Cereb Blood Flow Metab. 1981;1:413–7.

38. Willeumier KC, Taylor D V., Amen DG. Elevated BMI is associated with decreased blood flow in the prefrontal cortex using SPECT imaging in healthy adults. Obesity. 2011;19:1095–1097.

39. Knight SP, Laird E, Williamson W, et al. Obesity is associated with reduced cerebral blood flow – modified by physical activity. Neurobiol Aging. 2021;105:35–47.

40. Kullmann S, Heni M, Fritsche A, Preissl H. Insulin Action in the Human Brain: Evidence from Neuroimaging Studies. J Neuroendocrinol. 2015;27:419–423.

